# Auditory Complications among Childhood Cancer Survivors and Health-related Quality of Life: A PanCareLIFE study

**DOI:** 10.1101/2023.03.30.23286995

**Authors:** Sven Strebel, Katja Baust, Desiree Grabow, Julianne Byrne, Thorsten Langer, Antoinette am Zehnhoff-Dinnesen, Rahel Kuonen, Annette Weiss, Tomas Kepak, Jarmila Kruseova, Claire Berger, Gabriele Calaminus, Grit Sommer, Claudia E. Kuehni, the PanCareLIFE Consortium

## Abstract

Auditory complications are potential side effects from childhood cancer treatment. Yet, limited evidence exists about the impact of auditory complications—particularly tinnitus—on health-related quality of life (HRQoL) among childhood cancer survivors (CCS). We determined the prevalence of hearing loss and tinnitus in the large European PanCareLIFE cohort of CCS and examined its effect on HRQoL. We included CCS from four European countries who were diagnosed at age ≤ 18 years; survived ≥ 5 years; and aged 25–44 years at study. We assessed HRQoL (Short Form 36), hearing loss, and tinnitus using questionnaires. We used multivariable linear regression to examine associations between these two auditory complications and HRQoL adjusting for socio-demographic and clinical factors. Our study population consisted of 6,318 CCS (53% female; median age at cancer diagnosis 9 years interquartile range [IQR] 5–13 years) with median age at survey of 31 years (IQR 28–35 years). Prevalence was 7.5% (confidence interval [CI]: 6.9–8.2) for hearing loss and 7.6% (CI: 6.4–9.0) for tinnitus. CCS with hearing loss had impaired physical (coefficient [coef.] - 4.3, CI: -7.0 to -1.6) and mental (coef. -3.2, CI: -5.5 to -0.8) HRQoL when compared with CCS with normal hearing. Tinnitus was also associated with impaired physical (coef. -8.2, CI: -11.8 to -4.7) and mental (coef. -5.9, CI: -8.8 to -3.1) HRQoL. We observed the lowest HRQoL among CCS with both hearing loss and tinnitus. Our findings indicate timely treatment of hearing loss and tinnitus may contribute to quality of life of survivors.

## INTRODUCTION

Cancer treatment can cause auditory complications, such as hearing loss and tinnitus.^1, 2^ In recent surveys, childhood cancer survivors (CCS) reported more hearing loss and tinnitus when compared with their siblings.^3, 4^ Ototoxic cancer treatments include platinum-based chemotherapy, cranial radiotherapy (CRT), and surgeries involving the auditory system.^1, 4, 5^ Other suspected ototoxic treatments are concomitant medications such as aminoglycosides or loop diuretics, hematopoietic stem cell transplantation (HSCT), or the neurotoxic vinca alkaloid vincristine.^5-7^ Hearing loss and tinnitus lead to a wide range of educational and psychosocial problems such as learning difficulties and emotional distress among CCS and the general population.^8-11^ The overall burden of auditory complications ultimately affects health-related quality of life (HRQoL) of CCS.^12-14^

Only a few studies have investigated auditory complications and how they affect HRQoL among CCS.^12-14^ Previous studies with small sample sizes and heterogeneous inclusion criteria make comparisons between studies difficult since findings can only be extrapolated to the overall CCS population to a limited extent. The association of tinnitus with HRQoL among CCS remains unknown. Several studies examined the prevalence of hearing loss among CCS treated with cisplatin or CRT, yet studies of the overall population—and studies investigating tinnitus—are scarce.^3, 4, 15, 16^ We thus combined harmonized data from four European countries into a large cohort of CCS to describe the prevalence of hearing loss and tinnitus and investigate their association with HRQoL.

## METHODS

### Study population

PanCareLIFE (PCL) is a European-based study on late effects among CCS. It focuses on hearing loss, fertility problems, and quality of life.^17, 18^ For the current study, we included CCS from Switzerland (CH), Czech Republic (CZ), Germany (DE), and France (FR). The study population included national or regional cohorts of CCS 1) diagnosed with cancer according to the International Classification of Childhood Cancer (ICCC-3), 3rd edition,^19^ or Langerhans cell histiocytosis; 2) aged ≤18 years at time of cancer diagnosis; 3) survived ≥5 years after cancer diagnosis; 4) were off treatment for cancer at time of study; 5) aged 25-44 years when they received the questionnaire. To make data comparable between countries, we restricted our analysis to CCS ≥25 years and <45 years because data for CCS younger than 25 years were unavailable for the German cohort and data for CCS older than 45 years were unavailable for the French cohort. Details about study design, recruitment of participants, country-specific exclusion criteria, and characteristics of different cohorts were published in a separate study protocol.^17^

### Study procedure

Each country sent questionnaires to their respective regional or national cohorts between 2005 and 2017.^17^ The questionnaires were sent by mail except in CZ where clinic staff distributed them during follow-up visits to former patients. The questionnaire included questions about HRQoL, hearing, socio-demographic characteristics, and lifestyle behavior. Non-responders were reminded to complete the questionnaire.^17^ Clinical information on cancer diagnosis and treatment was extracted from medical records by each participating country.

### Assessment of HRQoL

We assessed HRQoL with the Short-Form 36 (SF-36) questionnaire.^20^ The SF-36 is a widely used instrument; several studies used it to determine HRQoL among CCS.^12, 21-24^ The questionnaire includes 36 items covering different aspects of physical and mental health aggregated into eight health domains: physical functioning (PF, 10 items), role-limitations due to physical problems (RP, 4 items), bodily pain (BP, 2 items), general health (GH, 5 items), vitality (VT, 4 items), social functioning (SF, 2 items), role-limitations due to emotional problems (RE, 3 items) and mental health (MH, 5 items).^20, 25^ These health domains are further collapsed into summary scores that reflect overall physical and mental health: physical component summary (PCS) and mental component summary (MCS). We converted all raw scores into T-scores ranging from 0–100 for each health domain. A higher score indicates better HRQoL. The T-scores were further transformed according to reference data from the German norm population stratified for age and sex (mean=50, SD=10).^17, 26^

### Auditory complications

We defined self-reported hearing loss (yes, no) and tinnitus (yes, no) as our main determinants of interest for impaired HRQoL. Participating country questionnaires contained slightly different worded questions on hearing (Supplement Table S1). The central PCL data center in Mainz (Germany) aggregated data and harmonized variables between participating countries in 2017.^17^ Data on tinnitus (yes, no) were unavailable for the German cohort; thus, we excluded German data for analyses involving tinnitus. We coded missing answers for hearing loss (<1%) and tinnitus (5%) as normal hearing and without tinnitus. We assumed that CCS with hearing loss or tinnitus would be more likely to answer the question than not when compared with CCS without auditory complications.

### Clinical and socio-demographic information

Based on previous study findings, we collected clinical and socio-demographic factors possibly associated with HRQoL among CCS: sex (female, male); age at survey; migration background (yes, no); education (primary, secondary, tertiary); occupational status (employed, unemployed); living with a partner (yes, no); currently smoking tobacco (yes, no); drinking >1 alcoholic beverage per week (yes, no); body mass index (BMI); cancer diagnosis according to ICCC-3^19^; age at diagnosis; history of relapse (yes, no); surgery (yes, no); chemotherapy (yes, no); radiotherapy (yes, no); HSCT (yes, no).^12, 21, 23, 27^ Respondents self-reported age at survey, migration background, education, occupational status, living with a partner, tobacco smoking status, alcohol consumption, and BMI variables.^17^ Demographic, cancer-related, and treatment information were extracted from participating institution medical records or corresponding cancer registries.^17^

### Statistical analysis

We used *t*-tests and fitted multivariable linear regression models to investigate possible associations of hearing loss or tinnitus with HRQoL. First, we examined whether mean scores on SF-36 health domains and PCS and MCS scores differed between CCS with and without auditory complications. We then fitted multivariable linear regression models to investigate whether any possible association of hearing loss or tinnitus with health domains and PCS and MCS scores were explained by clinical or socio-demographic factors. We chose linear regression because HRQoL outcome variables are continuous and binary categorizations of HRQoL measured by SF-36 is without consensus in the literature. To mitigate effects of sample imbalances between countries, we standardized cohorts from CZ, DE, and FR according to age at survey and sex variables. Because of the balanced distribution across all age groups and genders, we used the CH cohort as the reference population to calculate appropriate weights. Based on the conceptual framework of directed acyclic graphs (Supplement Figure S1),^28^ we adjusted our models for the following co-variables: age at survey (continuous in years); age at cancer diagnosis (continuous in years); type of cancer (categorical according ICCC-3); history of relapse (yes, no); surgery (yes, no); chemotherapy (yes, no); radiotherapy (yes, no); and HSCT (yes, no). We decided to include country of data provider to adjust for country-specific differences in recruitment of study participants and audiological monitoring.^17, 29, 30^ We calculated global *p*-values using the Wald test.

Since we hypothesized the burden of auditory complications may be largest when both tinnitus and hearing loss are present,^31^ we performed a sub-analysis to further investigate a potential causal relationship between auditory complications and HRQoL. We coded auditory complications as either 1) no auditory complications; 2) hearing loss only; 3) tinnitus only; or 4) hearing loss and tinnitus.

We used Stata version 16.1 (StataCorp LP, Austin, Texas) for all analyses.

## RESULTS

### Characteristics of study population

In total, 6,318 CCS were available for our analysis. Of the 6,318 CCS, most were from DE (n = 4,650; 74%); 822 (13%) from CH; 592 (9%) from CZ; and 254 (4%) from FR (Table 1). Our study population included 3,326 (53%) females and 2,992 (47%) males with median age of 31 (interquartile range [IQR] 28-35 years) at survey, median age 9 (IQR 5-13 years) at cancer diagnosis, and median 23 years (IQR 19-28) since cancer diagnosis. Leukemias (n = 2,033; 32%), lymphomas (n = 1,466; 23%), and central nervous system (CNS) tumors (n = 892; 14%) were most common cancer diagnoses. CCS received cancer treatment by surgery (2,580; 41%); chemotherapy (5,070; 80%); and radiotherapy (3,092; 49%).

**TABLE 1.**
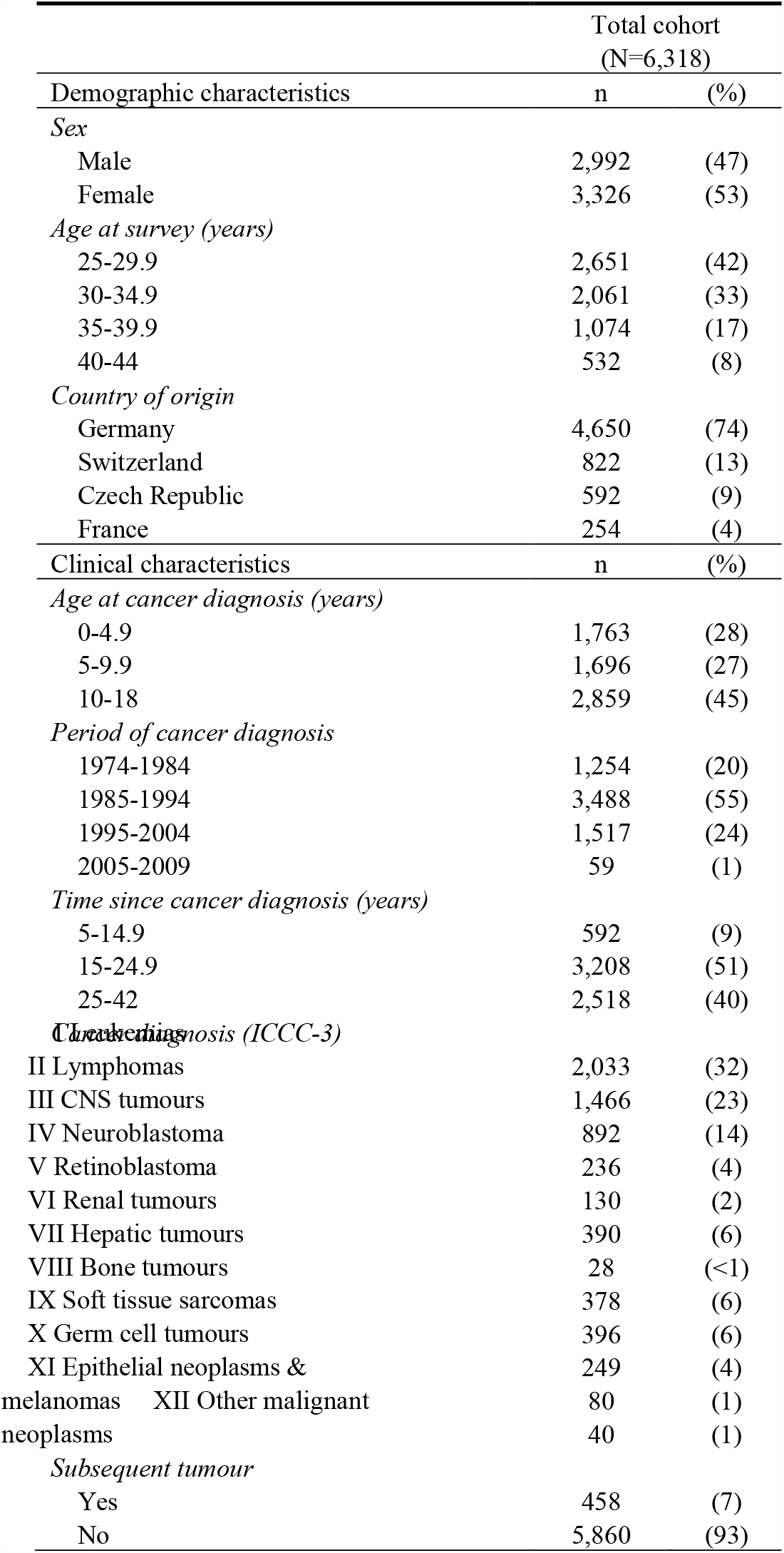
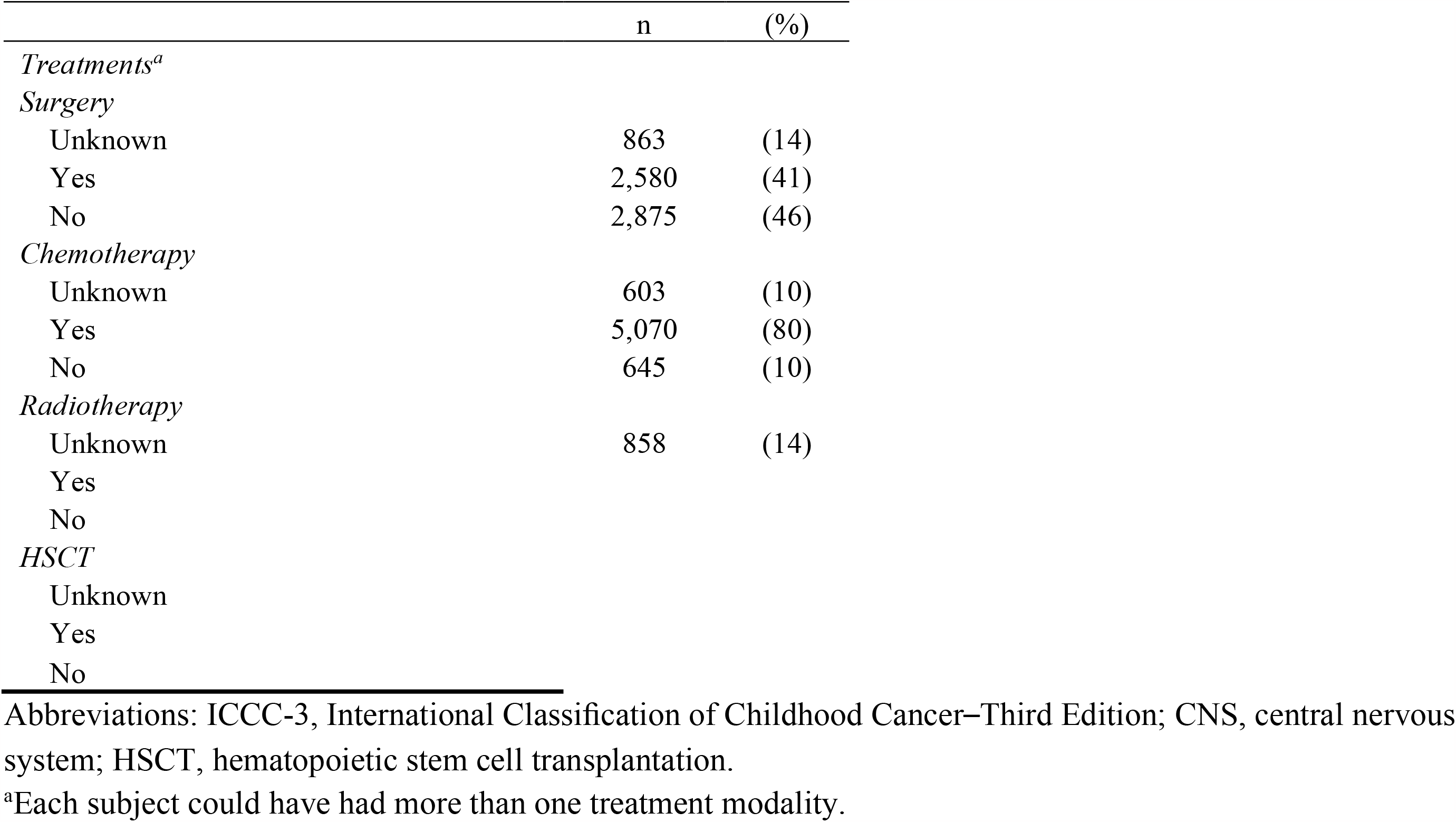
Demographic and clinical characteristics of study population

### Prevalence of auditory complications after childhood cancer

Of participating CCS, 7.5% (476/6,318; CI: 6.9-8.2) reported hearing loss. Data on tinnitus was available for the cohorts from CH, CZ, and FR resulting in a combined cohort of 1,668 CCS. Of those, 7.6% (127/1,668; CI: 6.4-9.0) reported tinnitus. Among CCS with tinnitus (n= 127), 45 (35%) also reported hearing loss. CCS diagnosed with CNS tumors, neuroblastoma, hepatic tumors, malignant bone tumors, soft tissue sarcomas, germ cell tumors, and epithelial neoplasms reported hearing loss more often than CCS diagnosed with leukemia (all *p* < 0.001) (Figure 1). CCS of hepatic tumors had the highest prevalence of hearing loss (8/28; 28.6%, CI: 13.2-48.7) followed by malignant bone tumors (91/378; 24.1%, CI: 19.8-28.7) and CNS tumors (130/892; 14.6%, CI: 12.3-17.1). Tinnitus prevalence was highest among CCS diagnosed with malignant bone tumors (16/103; 15.5%, CI: 9.1-24.0) and CNS tumors (33/255; 12.9%, CI: 9.1-17.7) (Figure 2).

**FIGURE 1.**
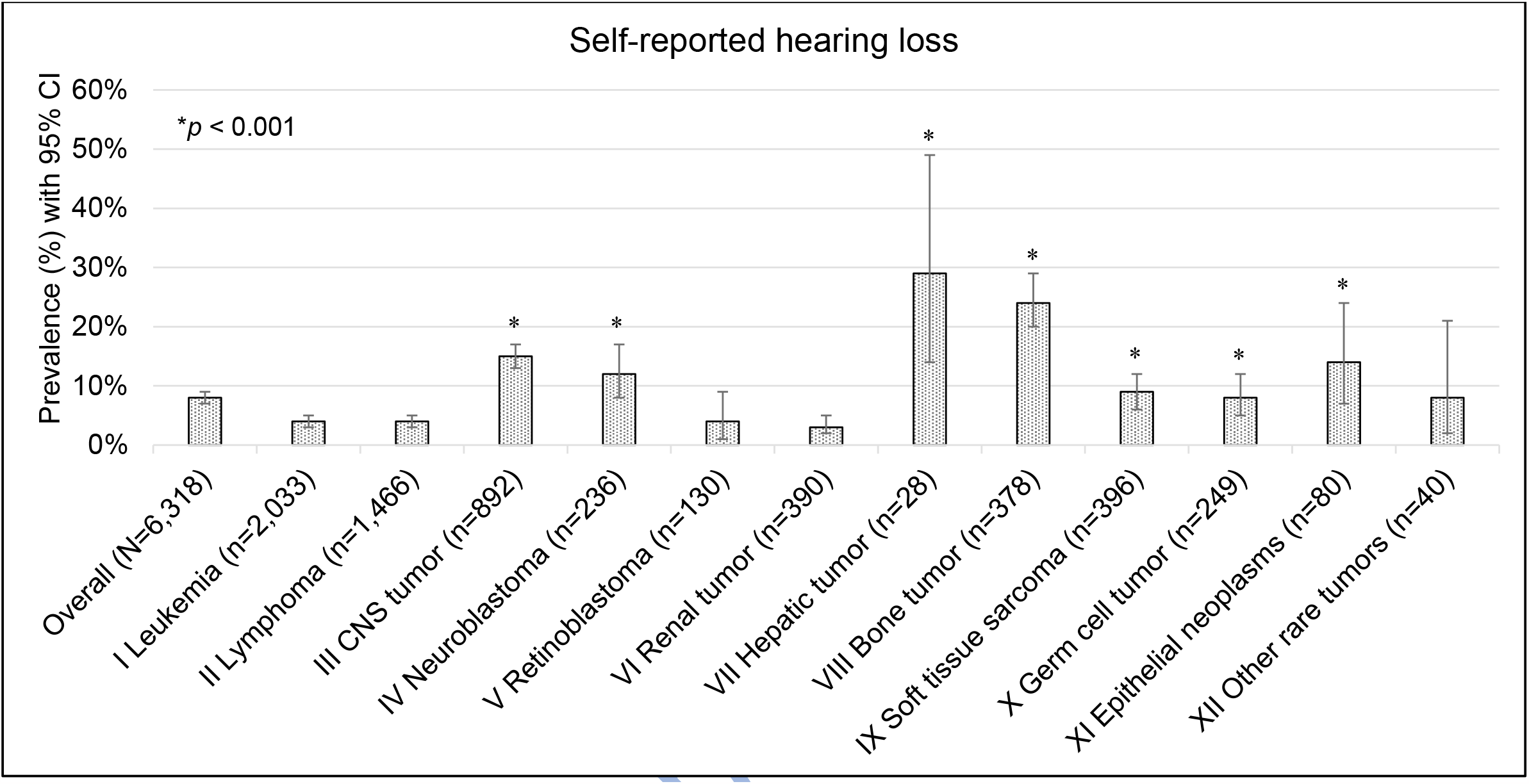
Prevalence of self-reported hearing loss at the time of the study (N=6,318). *P*-values are calculated from chi^2^-statistics comparing prevalence between survivors of leukemia with survivors of other tumor types. Abbreviations: CNS, central nervous system.

**FIGURE 2.**
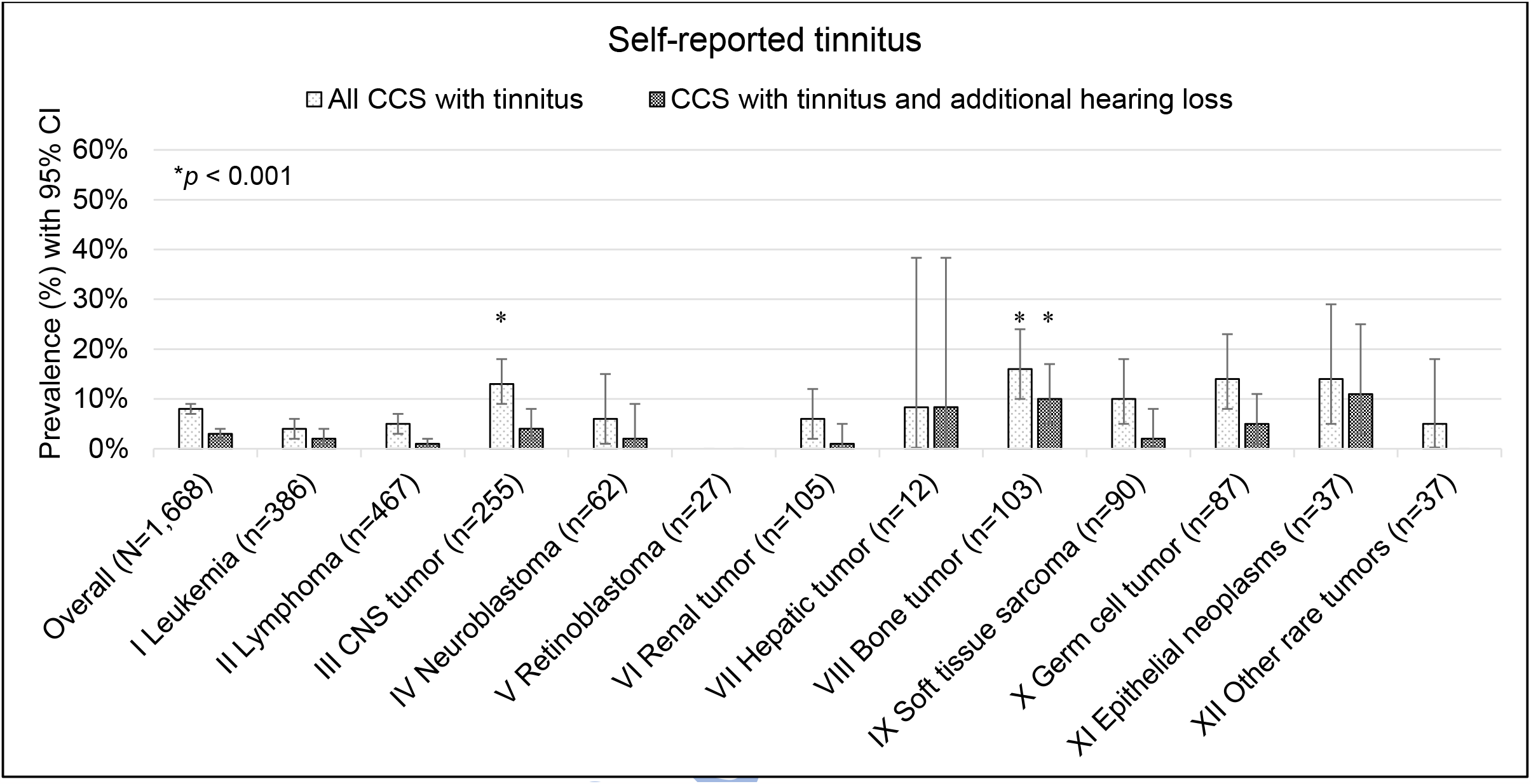
Prevalence of self-reported tinnitus at the time of the study (N=1,668). Data from the cohorts of CH, CZ, and FR are shown. No data on tinnitus was available for the German cohort (n=4,650). *P*-values are calculated from chi^2^-statistics comparing prevalence between survivors of leukemia with survivors of other tumor types. Abbreviations: CNS, central nervous system.

### Association of auditory complications with HRQoL

CCS with hearing loss had lower HRQoL mean scores than CCS with normal hearing (all differences with *p*<0.001) (Figure 3A). Looking at SF-36 summary scores, CCS with hearing loss scored 45.3 in overall physical (PCS) and 46.0 in overall mental (MCS) HRQoL. In comparison, CCS with normal hearing had 51.7 for PCS and 50.0 for MCS scores. Among the eight health domains, we observed the largest mean differences between CCS with and without hearing loss in physical functioning (40.6 vs. 48.3), general health (45.1 vs. 50.9), and social functioning (43.0 vs. 48.2).

**FIGURE 3.**
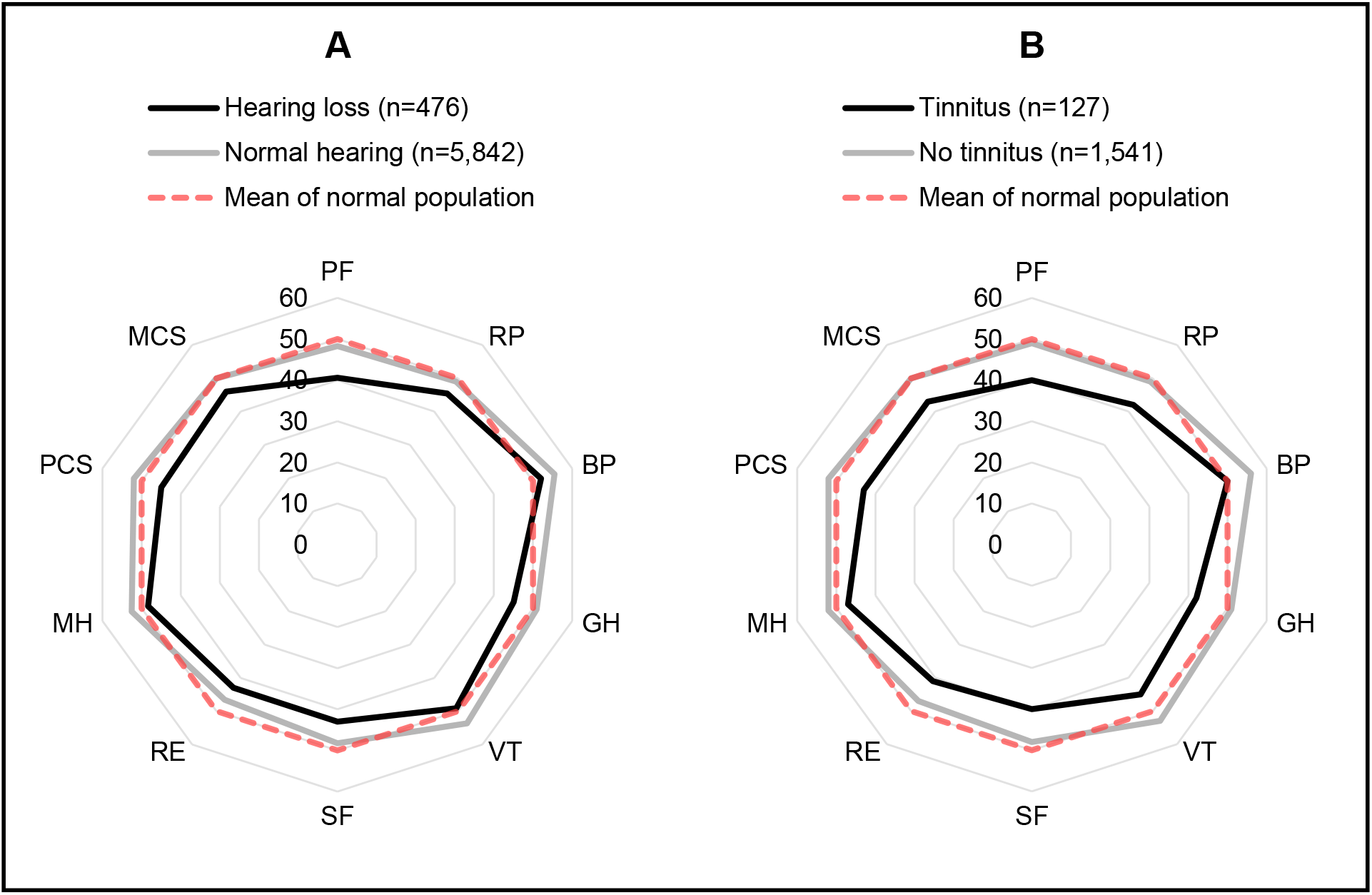
The two spider charts (A, B) show norm-based mean scores for all eight health domains and the two summary scores of the SF-36 comparing [A] CCS with hearing loss and normal hearing (N=6,318) and [B] CCS with tinnitus only, tinnitus and hearing loss, and without tinnitus (N=1,668). We included the cohorts from CH, CZ, and FR for the analysis of the association of tinnitus on HRQoL [B] (N=1,668) but excluded the cohort from Germany (n=4,650) because no data on tinnitus was available for the German cohort Higher scores indicate better HRQoL. Normal population (shown as dashed line) has an estimated mean score of 50 with a standard deviation of 10 for all HRQoL scores of the SF-36. Raw data of the figure are shown in the supplement (Supplement Table S2, S3). Abbreviations: PF, physical functioning; RP, role physical; BP, bodily pain; GH, general health; VT, vitality; SF, social functioning; RE, role emotional; MH, mental health; PCS, physical component summary; MCS, mental component summary.

CCS with tinnitus scored lower than CCS without tinnitus in all health domains and PCS (42.7 vs. 52.2) and MCS (43.1 vs. 49.9) summary scores (all differences with *p*<0.001) (Figure 3B). The largest mean differences between CCS with and without tinnitus were again in physical functioning (40.2 vs. 48.9), general health (41.6 vs. 51.0), and social functioning (39.7 vs. 48.3).

In the multivariable linear regression, hearing loss remained associated with lower HRQoL scores after adjusting for socio-demographic and cancer-related factors (Table 2). For PCS, coef. were -4.3 (CI: -7.0 to -1.6) among those with hearing loss and -3.2 (CI: -5.5 to -0.8) for MCS. On average, overall physical or mental HRQoL was reduced by 4.3 or 3.2 points for CCS with hearing loss compared with CCS with normal hearing. The association was strongest for general health, followed by physical functioning, vitality, and social functioning (coef. ranging from -4.6 to -3.8, *p*<0.05) (Table 2). We observed borderline or no associations of hearing loss in role physical and role emotional.

**TABLE 2.** Association of hearing loss and tinnitus with HRQoL from adjusted linear regression analysis

| Health Domains | Adjusted<sup>a</sup> association of hearing loss<sup>b</sup> |  | Adjusted<sup>a</sup> association of tinnitus<sup>c</sup> |  |
| --- | --- | --- | --- | --- |
|  | Coef. (95%-CI) | P<sup>d</sup> | Coef. (95%-CI) | P<sup>d</sup> |
| *Physical Functioning (PF)* | -4.5 (-7.7 to -1.4) | 0.005 | -7.2 (-11.2 to -3.2) | <0.001 |
| *Role Physical (RP)* | -2.3 (-4.6 to -0.1) | 0.044 | -5.9 (-8.8 to -2.9) | <0.001 |
| *Role Emotional (RE)* | -2.5 (-5.4 to 0.4) | 0.088 | -3.9 (-7.0 to -0.8) | 0.014 |
| *Bodily Pain (BP)* | -2.6 (-4.4 to -0.8) | 0.004 | -5.3 (-7.6 to -3.0) | <0.001 |
| *Mental Health (MH)* | -2.8 (-4.7 to -0.8) | 0.005 | -4.2 (-6.9 to -1.6) | 0.002 |
| *Vitality (VT)* | -3.9 (-6.0 to -1.8) | <0.001 | -8.4 (-11.3 to -5.6) | <0.001 |
| *General Health (GH)* | -4.6 (-6.9 to -2.3) | <0.001 | -8.3 (-11.4 to -5.3) | <0.001 |
| *Social Functioning (SF)* | -3.8 (-6.3 to -1.3) | 0.003 | -7.9 (-11.0 to -4.7) | <0.001 |
| Global Summary Scores | Coef. (95%-CI) | P<sup>d</sup> | Coef. (95%-CI) | P<sup>d</sup> |
| *PCS* | -4.3 (-7.0 to -1.6) | 0.002 | -8.2 (-11.8 to -4.7) | <0.001 |
| *MCS* | -3.2 (-5.5 to -0.8) | 0.008 | -5.9 (-8.8 to -3.1) | <0.001 |

Abbreviations: Coef., estimated beta coefficient from multivariable linear regression; PCS, physical component summary; MCS, mental component summary.
<sup>a</sup>Adjusted for: age at survey (continuous, in years); age at cancer diagnosis (continuous, in years); type of cancer diagnosis (according ICC-3); history of relapse (yes/no); surgery (yes, no); chemotherapy (yes, no); radiotherapy (yes, no); HSCT (yes, no); country of data provider.
<sup>b</sup>We included the total cohort for analysis of the association of hearing loss on HRQoL (N=6,318).
<sup>c</sup>We included the cohorts from CH, CZ, and FR for analysis of the association of tinnitus on HRQoL (N=1,668) but excluded the cohort from Germany (n=4,650) because no data on tinnitus was available for the German cohort.
<sup>d</sup>P-value calculated from Wald test.

Tinnitus also remained associated with lower HRQoL scores in multivariable linear regression (Table 2). The effects of tinnitus on PCS (coef. -8.2, CI: -11.8 to -4.7) and MCS (coef. -5.9, CI: -8.8 to -3.1) were greater compared with the effects of hearing loss (coef. of - 4.3 for PCS and -3.2 for MCS). We found the strongest effect of tinnitus on vitality (coef. - 8.4, CI: -11.3 to -5.6), general health (coef. -8.3, CI: -11.4 to -5.3), and social functioning (coef. -7.9, CI: -11.0 to -4.7) (all *p* < 0.001).

We found CCS with both tinnitus and hearing loss had lower overall physical and mental HRQoL compared with CCS with hearing loss alone (coef. -14.5 vs. -0.6 for PCS and coef. - vs. -2.9 for MCS) (Table 3). When compared with tinnitus alone, the effect of hearing loss and additional tinnitus was also larger for overall physical HRQoL (coef. -5.4 vs. -14.5 for PCS), yet similar for overall mental HRQoL (coef. -6.8 vs. -5.0 for MCS).

**TABLE 3.** Association of combined auditory complications on HRQoL from adjusted linear regression analysis

| Health Domains | Adjusted<sup>a</sup> association of<br>combined auditory complications |  |
| --- | --- | --- |
|  | Coef. (95%-CI) | *P*<sup>c</sup> |
| *Physical Functioning (PF)* |  | 0.002 |
| No auditory complications | *Reference* |  |
| Hearing loss only | -0.1 (-3.3 to 3.1) |  |
| Tinnitus only | -3.3 (-6.6 to 0.1) |  |
| Hearing loss and tinnitus | -15.5 (-24.8 to -6.3) |  |
| *Role Physical (RP)* |  | 0.002 |
| No auditory complications | *Reference* |  |
| Hearing loss only | 0.1 (-2.5 to 2.7) |  |
| Tinnitus only | -4.5 (-7.5 to -1.4) |  |
| Hearing loss and tinnitus | -8.8 (-15.2 to -2.5) |  |
| *Role Emotional (RE)* |  | 0.053 |
| No auditory complications | *Reference* |  |
| Hearing loss only | -2.0 (-6.2 to 2.2) |  |
| Tinnitus only | -4.0 (-7.2 to -0.8) |  |
| Hearing loss and tinnitus | -4.3 (-11.0 to 2.3) |  |
| *Bodily Pain (BP)* |  | <0.001 |
| No auditory complications | *Reference* |  |
| Hearing loss only | -1.1 (-3.5 to 1.2) |  |
| Tinnitus only | -4.4 (-7.0 to -1.7) |  |
| Hearing loss and tinnitus | -7.6 (-11.7 to -3.5) |  |
| *Mental Health (MH)* |  | 0.007 |
| No auditory complications | *Reference* |  |
| Hearing loss only | -2.3 (-5.1 to 0.6) |  |
| Tinnitus only | -4.5 (-7.8 to -1.1) |  |
| Hearing loss and tinnitus | -4.3 (-8.3 to -0.3) |  |
| *Vitality (VT)* |  | <0.001 |
| No auditory complications | *Reference* |  |
| Hearing loss only | -3.0 (-6.1 to 0.1) |  |
| Tinnitus only | -8.7 (-12.2 to -5.2) |  |
| Hearing loss and tinnitus | -8.6 (-13.1 to -4.2) |  |

**TABLE 3** Continued.
| Health Domains | Coef. (95%-CI) | *P*<sup>c</sup> |
| --- | --- | --- |
| *General Health (GH)* |  | <0.001 |
| No auditory complications | *Reference* |  |
| Hearing loss only | -2.6 (-6.1 to 0.9) |  |
| Tinnitus only | -7.2 (-11.0 to -3.4) |  |
| Hearing loss and tinnitus | -11.4 (-15.8 to -7.0) |  |
| *Social Functioning (SF)* |  | <0.001 |
| No auditory complications | *Reference* |  |
| Hearing loss only | -1.6 (-5.0 to 1.8) |  |
| Tinnitus only | -7.0 (-10.4 to -3.6) |  |
| Hearing loss and tinnitus | -10.0 (-16.2 to -3.9) |  |
| Global Summary Scores | Coef. (95%-CI) | *P*<sup>c</sup> |
| *PCS* |  | <0.001 |
| No auditory complications | *Reference* |  |
| Hearing loss only | -0.6 (-3.5 to 2.2) |  |
| Tinnitus only | -5.4 (-8.8 to -1.9) |  |
| Hearing loss and tinnitus | -14.5 (-21.9 to -7.1) |  |
| *MCS* |  | <0.001 |
| No auditory complications | *Reference* |  |
| Hearing loss only | -2.9 (-6.2 to 0.5) |  |
| Tinnitus only | -6.8 (-10.1 to -3.5) |  |
| Hearing loss and tinnitus | -5.0 (-10.0 to 0.1) |  |

Abbreviations: PCS, physical component summary; MCS, mental component summary.
<sup>a</sup>Adjusted for: age at survey (continuous, in years); age at cancer diagnosis (continuous, in years); type of cancer diagnosis (according ICCC-3); history of relapse (yes/no); surgery (yes, no); chemotherapy (yes, no); radiotherapy (yes, no); HSCT (yes, no); country of data provider.
<sup>b</sup>We included the cohorts from CH, CZ, and FR for the analysis (N=1,668) but excluded the cohort from Germany (n=4,650) because no data on tinnitus was available for the German cohort.
<sup>c</sup>*P*-value calculated from Wald test.

## DISCUSSION

We found the prevalence of auditory complications varied between cancer diagnoses and the highest prevalence of hearing loss and tinnitus among survivors of CNS and malignant bone tumors. HRQoL was lower among CCS with auditory complications than for those with normal hearing. Hearing loss and tinnitus were strongly associated with physical functioning, vitality, general health, and social functioning. We observed lower HRQoL among CCS with hearing loss and additional tinnitus compared with CCS with hearing loss alone.

### Strengths and limitations

Since our study is the largest cohort of CCS to examine auditory complications and their association with HRQoL, it results in high statistical power and good representativeness because it combined data from population-based and regional well-defined cohorts. Tinnitus is more frequent among CCS compared with the general population,^2^ yet its association with HRQoL among CCS was unknown. We included CCS with all possible cancer treatments, not only those exposed to ototoxic treatments such as platinum-based chemotherapy or CRT, which allowed assessing the burden of auditory complications among the overall population of CCS.^32^ We applied SF-36—an established and validated instrument widely used in previous studies—to measure HRQoL among CCS, which allows comparing our data with other studies.^21-23, 27, 33, 34^ Data from participating countries were collected centrally and harmonized before merging to avoid data management errors.^17^ However, our study results might still be influenced by study design differences of participating countries, leading to potential selection bias. For example, FR did not contact CCS of leukemias, and—similar to DE—sent questionnaires later (≥10 years) than CH and CZ (both ≥5 years) after cancer diagnosis.^17^ Considering the French cohort represents only 4% of the total study population, we assume that selection based on cancer diagnosis did not result in a major bias in our findings. Additionally, time since diagnosis was investigated in two larger population-based studies showing either no or minor effects on HRQoL among CCS.^23, 27^ Other limitations relate to the main exposures of interest; hearing loss and tinnitus. Since auditory complications were assessed by questionnaire and dependent on severity, underreporting is possible. For instance, CCS with severe hearing loss possibly received better audiologic care and recall it better than CCS with mild high-frequency hearing loss who are unaware of it.^35^ Additionally, SF-36 does not specifically measure HRQoL related to hearing and may not capture life situations affected by auditory complications.

### Comparison with previous studies

Among our study population, 7.5% of CCS reported hearing loss. We observed particularly high prevalence among survivors of hepatoblastoma, CNS tumors, and malignant bone tumors—an expected finding from higher cisplatin or CRT use compared with other cancer treatment regimes.^5, 7^ Larger studies on hearing loss prevalence among CCS mostly focused on high-risk populations treated with platinum-based chemotherapy or CRT.^5-7^ Two population-based studies from Switzerland (Swiss Childhood Cancer Survivor Study; SCCSS) and the United States (Childhood Cancer Survivor Study; CCSS) determined the prevalence of hearing loss among the overall CCS population with questionnaires.^3, 16^ The Swiss population in our cohort overlaps with the study population of the SCCSS.^3, 17, 36^ Therefore, we only compared our data with the CCSS study.^16^ Whelan and colleagues found a prevalence of self-reported hearing loss of 5%, which is slightly lower than what we found (7.5%).^16^ They included CCS diagnosed in earlier years (1970-1986) compared with our study (1974-2009), which possibly explains the difference. Considering cisplatin was first approved in 1978 for adult cancer treatment, it is possible a higher proportion of CCS in our cohort were treated with ototoxic platinum-based chemotherapy, as Weiss and colleagues also discuss in their SCCSS study.^3, 37^

The prevalence of tinnitus was 7.6% among our study population. CCS diagnosed with CNS tumors or malignant bone tumors had a three to four times higher prevalence compared with survivors of leukemias. The higher prevalence is possibly explained by previously identified risk factors for tinnitus among CCS, such as exposure to cisplatin, CRT, and CNS surgeries.^4^ In Meijer and colleagues’ systematic review, the prevalence of tinnitus ranged from 3-17%.^2^ They also recently published a population-based study where they estimated the prevalence of tinnitus to be 9.5% among CCS compared with 3.7% for siblings.^4^ Their findings are consistent with our study.

Audiological complications were associated with lower HRQoL, particularly with decreased physical functioning, general health, vitality, and social functioning. Physical functioning reflects limitations in physical activities, such as difficulties walking a mile or exercising vigorously due to health problems.^20^ In a SCCSS study, physical well-being was lower among younger CCS with hearing loss than for CCS with normal hearing.^12^ General health reflects current and future health perceptions; for example, how people perceive their health when compared with peers or whether their health deteriorates in the future.^20^ General health was also heavily impaired among CCS when compared with siblings or the general population in previous studies.^23, 27^ The SF-36 assesses vitality with questions such as whether people feel full of energy or tired and worn out.^20^ Previous studies of the general population showed—depending on severity—patients with tinnitus experience comorbidities, such as sleep disturbance, fatigue, and depression.^10, 11, 38^ Hearing loss possibly leads to feelings of fatigue from long periods of effortful listening.^39-41^ Impaired social functioning refers to limitations in social activities, such as visiting family and friends, due to physical or emotional health problems.^20^ CCS with hearing loss reported psychosocial difficulties and communication problems in previous studies examining the impact of hearing loss on HRQoL.^12-14^ Yet, a direct comparison with our study remains difficult because they only included children and adolescents—a study population whose social behavior differs from our adult study population (median age 31 at survey). Data from adult CCS participating in the St. Jude Lifetime Cohort Study showed treatment-related hearing loss associated with reduced social attainment, which possibly relates to decreased social engagement.^42^ However, none of these studies investigated the impact of tinnitus on social behavior and attainment.

### Causality between auditory complications and HRQoL

The observed association of lower vitality and social functioning possibly relates to educational and psychosocial problems caused by auditory complications.^8, 10, 11, 13^ Other chronic health problems, such as musculoskeletal or neurological, also affect HRQoL.^27^ In our study, we could not adjust for other chronic health problems. Since the risk of auditory complications and other chronic health problems increases with more intensive cancer treatment, unobserved late effects in other organ systems could contribute to lower physical and mental HRQoL (Supplement Figure S1).^4, 5, 7, 43, 44^ However, we observed hearing loss with additional tinnitus reduces HRQoL more than hearing loss alone. Since we assumed the burden on daily life is greater when CCS experience both hearing loss and tinnitus, it possibly indicates a causal relationship.^31^ Interestingly, tinnitus alone also had a greater impact on HRQoL than hearing loss alone. Since data are self-reported and tinnitus is probably underreported in our study, further research using objective hearing tests and validated instruments to assess tinnitus are important to understand its impact on CCS.^30, 45^

### Conclusion

Our study showed hearing loss and tinnitus strongly affected HRQoL among CCS— particularly reduced HRQoL among survivors with both, hearing loss and tinnitus. Our findings support current guideline recommendations for timely referrals to audiologists for tinnitus symptoms and optimized treatment of hearing loss and tinnitus since both affect HRQoL.^30^

## Data Availability

The data that support the information of this manuscript were accessed on secured servers of the Institute of Social and Preventive Medicine at the University of Bern. Individual-level sensitive data can only be made available for researchers who fulfil the respective legal requirements. All data requests should be communicated to the corresponding author.

## Abbreviations

BMI: Body Mass Index
BP: Bodily pain
CCS: Childhood cancer survivors
CCSS: American Childhood Cancer Survivor Study
CH: Switzerland
CI 95%: Confidence interval
CNS: Central nervous system
Coef.: Estimated beta coefficient from linear regression
CRT: Cranial radiation therapy
CZ: The Czech Republic
DE: Germany
FR: France
GH: General health
HRQoL: Health-related quality of life
HSCT: hematopoietic stem cell transplantation
ICCC-3: International Classification of Childhood Cancer, Third edition
IQR: Interquartile range
MCS: Mental component summary
MH: Mental health
PCL: PanCareLIFE
PCS: Physical component summary
PF: Physical functioning
RE: Role emotional
RP: Role physical
SCCSS: Swiss Childhood Cancer Survivor Study
SF: Social functioning
SF-36: Short-Form 36 questionnaire
VT: Vitality

## AUTHORS CONTRIBUTIONS

SST and KB are the first authors of this paper and both contributed substantially to this work. SST prepared the final data set, wrote the manuscript, performed the data analysis and interpretated the results. KB coordinated the study, contributed to the design, implementation, data collection and harmonization, data preparation, reviewed the manuscript, and commented on data analysis and interpretation. DG and JB were part of the PanCareLIFE coordination team and contributed to the implementation and coordination of the study; they also supervised collection, and harmonization and maintenance of data. TL and AaZD contributed to the study implementation and to data harmonization and they provided their medical expertise for data analysis and interpretation. RK, AW, TK, JK, CB conducted the study in the different countries and contributed to data collection and harmonization. AW was involved in the study design. GC supervised the design, implementation and coordination of the study, and contributed to data collection, harmonization and analysis. GS and CK are senior authors of the study; they were involved in the study design and implementation, in collection, harmonization, and maintenance of data, they supervised analysis and interpretation of data and reviewed the manuscript. All co-authors provided feedback on the manuscript and approved its final version.

## ACKNOWLEDGMENTS

We thank all survivors of childhood and adolescent cancers and their families for their participation in our study. PanCareLIFE (Grant no. 602030) is a collaborative project in the 7th Framework Program of the European Union. Project partners are Universitätsmedizin der Johannes Gutenberg-Universität Mainz, Germany (PD Dr P Kaatsch, Dr D Grabow); Boyne Research Institute, Drogheda, Ireland (Dr J Byrne, Ms H Campbell); Pintail Ltd., Dublin, Ireland (Mr C Clissmann, Dr K O’Brien); Academisch Medisch Centrum bij de Universiteit van Amsterdam, the Netherlands (Prof Dr LCM Kremer); Universität zu Lübeck, Germany (Prof T Langer); Stichting VU-VUMC, Amsterdam, the Netherlands (Dr E van Dulmen-den Broeder, Dr MH van den Berg); Erasmus Universitair Medisch Centrum, Rotterdam, the Netherlands; and Princess Maxima Center for Pediatric Oncology, Utrecht, the Netherlands (Prof Dr MM van den Heuvel-Eibrink); Charité-Universitätsmedizin Berlin, Germany (Prof Dr A Borgmann-Staudt); Westfälische Wilhelms-Universität Münster, Germany (Prof Dr A am Zehnhoff-Dinnesen); Universität Bern, Switzerland (Prof Dr CE Kuehni); Istituto Giannina Gaslini, Genoa, Italy (Dr R Haupt); Fakultni nemocnice Brno, Czech Republic (Dr T Kepak); Centre Hospitalier Universitaire Saint-Étienne, Saint-Étienne, France (Dr C Berger); Kraeftens Bekaempelse, Copenhagen, Denmark (Dr JF Winther); Fakultni nemocnice v Motole, Prague, the Czech Republic (Dr J Kruseova); Universitaetsklinikum Bonn, Bonn, Germany (Dr G Calaminus, K Baust); and University Hospital Essen, Essen, Germany (Prof U Dirksen). The EU Commission takes no responsibility for any use made of the information set out.

In Switzerland, we thank the Swiss Childhood Cancer Survivor Study team, the Swiss Paediatric Oncology Group data managers, and the Swiss Childhood Cancer Registry study team. We also thank Marcel Zwahlen for statistical advice. We thank the editorial service of the Institute of Social and Preventive Medicine at the University of Bern for the editorial suggestions.

In Czech Republic, at the University Hospital of Brno we thank the team of collaborators from the Children’s Hospital in Brno, Masaryk University in Brno, especially Katerina Kepakova, and Brno survivors’ association Together Toward a Smile: H Hrstková, V BajČiová, D Hošnová, Z Kuttnerová, M Blanářová, R Mazúr, I Mikulec, E BuČková, L Červinková, E Bařinová, I Krupková, E Novotná, P Chloupková, Z Wimmerová, L Štrublová, and many other external collaborators and supporters.

At the University Hospital of Prague LTFU Care Registry, the authors thank the team of the Prague Childhood Cancer Survivor Study: Aleš Lukš, P Keslová, M Ganevová, V Reichlová, J Bašeová, L Nováková, M Douchová, and M Čepelová.

In France, we thank the team of the long-term follow-up study (SALTO): Faure-Conter (Lyon), N Corradini (Lyon), D Plantaz (Grenoble), E Tarral (Grenoble), I Durieu (Lyon), I Guichard (Saint-Étienne), L Casagranda and F Odier (Saint-Étienne), N Gauthier (Lyon), P Métral (Lyon), M Mercier (Lyon), S Billet (Grenoble), C Celette (Grenoble), I Schiff (Grenoble), and A Loubier (Saint-Étienne).

In Germany, we thank the team of the Clinic of Phoniatrics and Pedaudiology at the University Hospital in Münster and theVIVE group and members of the GCCR: U Creutzig (Hannover), T Langer (Lübeck), J Dobke (Berlin), I Jung (Mainz), I Kerenyi (Mainz), C Teske (Bonn), and M Zimmermann (Hannover). In Germany, they also thank the team at GCCR in its role as central data center for PanCareLIFE (Peter Kaatsch, Claudia Spix, Melanie Kaiser, Claudia Bremensdorfer).

Our project received funding from the European Union’s Seventh Framework Program for research, technological development, and demonstration under Grant no. 602030.

In Switzerland, our project received funding from the Swiss Cancer League (Grant no. KLS-3412-02-2014, KLS-3886-02-2016, and HSR-4951-11-2019), the Swiss Cancer Research Foundation (Grant no. KFS-4157-02-2017), the Bernese Cancer League, Kinderkrebshilfe Schweiz, Kinderkrebs Schweiz, and the CANSEARCH Foundation (https://cansearch.ch/en/). The work of the SCCR was supported by the Swiss Pediatric Oncology Group, Schweizerische Konferenz der kantonalen Gesundheitsdirektorinnen und - direktoren, the Swiss Cancer Research, Kinderkrebshilfe Schweiz, Bundesamt für Gesundheit, National Institute for Epidemiology and Cancer Registration, and Celgene.

In the Czech Republic, our study received funding from the University Hospital of Brno from the Ministry of Education, Youth and Sports of the Czech Republic, under Grant no. 7E13061 and the University Hospital of Prague from the foundation Národ dětem.

In France, our study received funding from The Wyeth Foundation, the French National Institute of Cancer (INCa), and the French League against Cancer.

In Germany, our study received funding from the German Cancer Aid (Grant No. 110298). Data for this subproject were provided by University of Bern, Switzerland (Prof CE Kuehni); Fakultni nemocnice Brno, Czech Republic (Dr T Kepak); Centre Hospitalier Universitaire Saint-Étienne, France (Dr C Berger); Fakultni nemocnice v Motole, Prague, Czech Republic (Dr J Kruseova); and Universitaetsklinikum Bonn, Bonn, Germany (Dr G Calaminus).

Prof Dr NW Paul at the University of Mainz and Prof Dr Lisbeth E Knudsen from the University of Copenhagen provided independent ethics advice.

## ETHICS STATEMENT

PanCareLIFE is a multinational collaborative research project that has harmonized and combined data from regional and national cohorts across Europe. Data collection and analysis for each cohort was approved by the responsible ethics committee in each participating country. For Germany this is the Ethics Committee of the Medical Association of Westphalia-Lippe and the Medical faculty of the Westphalian Wilhelms University (2012-530-f-S), for Switzerland the Cantonal Ethics Committee of the Canton of Bern (KEK-BE: 166/2014; 2021-01462), for the Czech Republic the Ethics Committee for Multi-Centric Clinical Trials of the University Hospital Motol (EK-1723/13) and the Multi-Centric Ethics Committee of the University Hospital Brno (approval date: 2014/10/22), and for France the Personal Protection Committee South East 1 (CPP: 2015-23).

## LEGENDS

**Supplementary Table S1** Questions on auditory complications from the questionnaires of each participating country translated into English

**Supplementary Table S2** Norm-based mean scores from SF-36 comparing CCS with hearing loss and normal hearing

**Supplementary Table S3** Norm-based mean scores from SF-36 comparing CCS with and without tinnitus

**Supplementary Figure S1** Directed acyclic graph (DAG)

