## Supplementary information for "Auditory Complications among Childhood Cancer Survivors and Health-related Quality of Life: A PanCareLIFE study"

**Supplementary data**

**SUPPLEMENTARY TABLE S1** Questions on auditory complications from the questionnaires of each participating country translated into English.

| Country | Corresponding variable | Question |
| --- | --- | --- |
| Switzerland | Hearing loss (yes, no) | Have you ever been told by a doctor that you have, or have had problems with hearing? |
| | Tinnitus (yes, no) | Have you ever been told by a doctor that you have, or have had problems with tinnitus (ringing in the ears)? |
| The Czech Republic | Hearing loss (yes, no) | Have you been informed by a doctor that you suffer from hearing impairment? |
| | Tinnitus (yes, no) | Have you been informed by a doctor that you suffer from tinnitus? |
| France | Hearing loss (yes, no) | Has a doctor, nurse, or other health care professional told you that you have or have had problems with hearing? |
| | Tinnitus (yes, no) | Do you have or have you ever had tinnitus or ringing in the ears? |
| Germany[a] | Hearing loss (yes, no) | Have you been diagnosed with a hearing loss? |

[a]No data on tinnitus were available for the German cohort.

non peer-reviewed

**SUPPLEMENTARY TABLE S2** Norm-based[a] mean scores from SF-36 comparing CCS with hearing loss and normal hearing

| | All combined N=6,318 | | |
|---|---|---|---|
| Health Domains | Mean | (95% CI) | $P^b$ |
| *Physical Functioning (PF)* | | | <0.001 |
| Hearing loss | 40.6 | (38.9-42.3) | |
| Normal hearing | 48.3 | (47.9-48.6) | |
| *Role Physical (RP)* | | | <0.001 |
| Hearing loss | 45.4 | (44.1-46.7) | |
| Normal hearing | 49.0 | (48.8-49.3) | |
| *Bodily Pain (BP)* | | | <0.001 |
| Hearing loss | 52.0 | (51.1-53.0) | |
| Normal hearing | 55.5 | (55.2-55.7) | |
| *General Health (GH)* | | | <0.001 |
| Hearing loss | 45.1 | (43.7-46.4) | |
| Normal hearing | 50.9 | (50.5-51.2) | |
| *Vitality (VT)* | | | <0.001 |
| Hearing loss | 49.1 | (47.9-50.3) | |
| Normal hearing | 53.6 | (53.3-53.9) | |
| *Social functioning (SF)* | | | <0.001 |
| Hearing loss | 43.0 | (41.7-44.3) | |
| Normal hearing | 48.2 | (47.9-48.6) | |
| *Role Emotional (RE)* | | | <0.001 |
| Hearing loss | 42.9 | (41.4-44.5) | |
| Normal hearing | 46.6 | (46.2-46.9) | |
| *Mental Health (MH)* | | | <0.001 |
| Hearing loss | 48.3 | (47.2-49.5) | |
| Normal hearing | 52.5 | (52.2-52.8) | |
| Global Summary Scores | Mean | (95% CI) | $P^b$ |
| *PCS* | | | <0.001 |
| Hearing loss | 45.3 | (43.8-46.7) | |
| Normal hearing | 51.7 | (51.4-52.0) | |
| *MCS* | | | <0.001 |
| Hearing loss | 46.0 | (44.7-47.4) | |
| Normal hearing | 50.0 | (49.7-50.4) | |

Abbreviations: SF-36, Short-Form 36 questionnaire; CCS, childhood cancer survivors; CNS, central nervous system; CI, Confidence interval; PCS, physical component summary score; MCS, mental component summary score

[a]Norm-based to the German normal population with a mean of 50 and standard deviation of 10.

[b]*P*-values calculated from two-tailed t-tests comparing CCS with hearing loss and with normal hearing.

non peer-reviewed

**SUPPLEMENTARY TABLE S3** Norm-based[a] mean scores from SF-36 comparing CCS with and without tinnitus

| | All combined (N=1,668) | | |
|---|---|---|---|
| Health Domains | Mean | (95% CI) | $P^{c}$ |
| *Physical Functioning (PF)* | | | <0.001 |
| Hearing loss | 40.2 | (36.8-43.5) | |
| Normal hearing | 48.9 | (48.3-49.5) | |
| *Role Physical (RP)* | | | <0.001 |
| Hearing loss | 42.2 | (39.8-44.7) | |
| Normal hearing | 49.4 | (48.9-49.9) | |
| *Bodily Pain (BP)* | | | <0.001 |
| Hearing loss | 49.8 | (47.8-51.7) | |
| Normal hearing | 55.9 | (55.4-56.3) | |
| *General Health (GH)* | | | <0.001 |
| Hearing loss | 41.6 | (39.0-44.3) | |
| Normal hearing | 51.0 | (50.3-51.7) | |
| *Vitality (VT)* | | | <0.001 |
| Hearing loss | 44.9 | (42.5-47.4) | |
| Normal hearing | 53.4 | (52.7-54.0) | |
| *Social functioning (SF)* | | | <0.001 |
| Hearing loss | 39.7 | (36.9-42.4) | |
| Normal hearing | 48.3 | (47.7-48.9) | |
| *Role Emotional (RE)* | | | <0.001 |
| Hearing loss | 41.5 | (38.8-44.2) | |
| Normal hearing | 47.1 | (46.5-47.7) | |
| *Mental Health (MH)* | | | <0.001 |
| Hearing loss | 46.9 | (44.6-49.2) | |
| Normal hearing | 52.2 | (51.6-52.7) | |
| Global Summary Scores | Mean | (95% CI) | $P^{c}$ |
| *PCS* | | | <0.001 |
| Hearing loss | 42.7 | (39.7-45.6) | |
| Normal hearing | 52.2 | (51.6-52.8) | |
| *MCS* | | | <0.001 |
| Hearing loss | 43.1 | (40.4-45.7) | |
| Normal hearing | 49.9 | (49.2-50.5) | |

Abbreviations: SF-36, Short-Form 36 questionnaire; CCS, childhood cancer survivors; CNS, central nervous system; CI, Confidence interval; PCS, physical component summary score; MCS, mental component summary score

[a]Norm-based to the German normal population with a mean of 50 and standard deviation of 10.

[b]Data on tinnitus (yes or no) were not available for the German cohort (n=4,627).

[c]*P*-values calculated from two-tailed t-tests comparing CCS with and without tinnitus.

non peer-reviewed

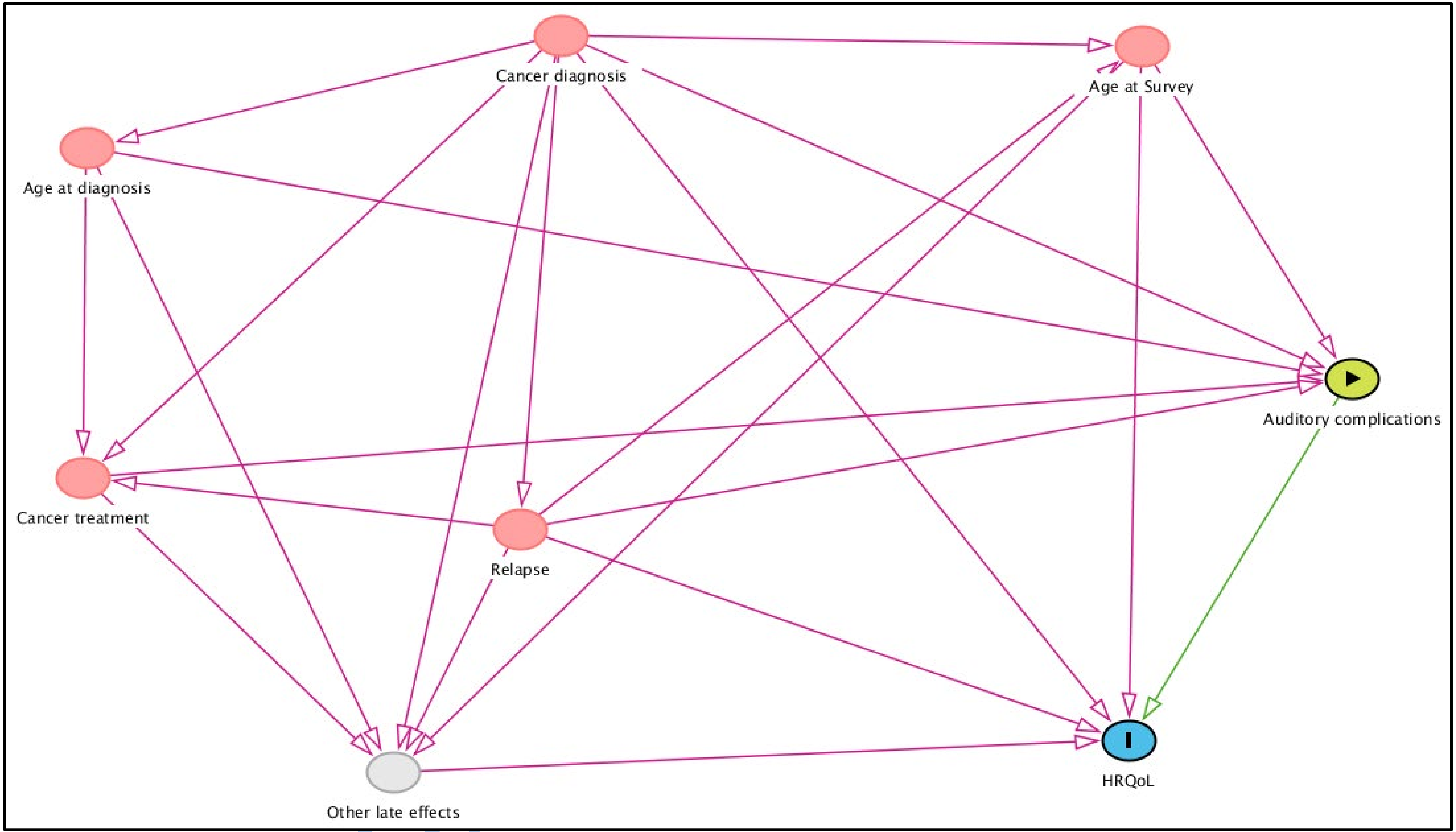


**SUPPLEMENTARY FIGURE S1** The directed acyclic graph (DAG) shows factors that may be associated with auditory complications and HRQoL but are not on the presumed causal pathway between auditory complications and impaired HRQoL. We did not collect data on other late effects. For a better understanding of the associations in this DAG, we added it as a non-observed variable. For all of these variables except the non-observed other late effects, we adjusted our multivariable linear regression model.
